# Hospital Standardised Length of Stay Ratio

**DOI:** 10.1101/2025.06.26.25330250

**Authors:** Graeme J Duke, Steven Hirth, John D Santamaria, Zhuoyang Li, Carla Read, Adina Hamilton, Elsa Lapiz, Teresa Le, Tharanga Fernando, Rosangela Merlo

## Abstract

**Objective:** Hospital length of stay (LOS) is a key indicator of hospital efficiency and quality of care, but a reliable metric for benchmarking LOS remains problematic. This report describes the methodology to generate a standardised hospital LOS ratio (HSLR).

**Design:** Retrospective observational analysis of LOS from an administrative dataset using a survival (time-to-event) analytic approach to generate average (aLOS) and risk-adjusted LOS (pLOS), and the HSLR (= [sum observed LOS]/[sum total pLOS]).

**Setting:** 334 (public and private) hospitals in the state of Victoria, Australia, adult population 5.28 million.

**Participants:** 2.73 million adult multi-day acute-care hospital separations and 15.53 million bed-days over five years, July 2019 - June 2024.

**Main outcomes:** Hospital aLOS, pLOS, and HSLR at the provider level with model fit assessed for calibration (Cox-Snell residuals), classification (aLOS and HSLR results for each hospital-years compared to benchmark), variance (intra-class correlation coefficient [ICC]) and dispersion (value [ϕ] and random effect standard deviation [τ]) characteristics.

**Results:** LOS was markedly right skewed and autocorrelated (p<0.001); population median 3.94 (interquartile range: 2.48-6.76); aLOS 5.68 ±4.98 days. LOS prediction model included six demographic covariates (age, sex, aged-care residency, emergent, admission source, unplanned transfer) and 12,145 separate principal diagnoses aggregated into nine ranked LOS risk-categories. 572 (61% of 940 hospital-year) aLOS values were outside ±3SD benchmark; whereas 936 (99.5%) HSLR values within ±3SD; 98% within ±2SD. Model dispersion (ϕ = 2.80; τ = 0.15) and ICC at provider level (0.025) were low.

**Conclusions:** aLOS is a simple descriptor but poor comparator. Time-to-event survival analytic models furnish risk-adjusted pLOS and HSLR metrics which indicate that majority of LOS variation is due to patient-related, not hospital, factors.

## INTRODUCTION

Hospital length of stay (LOS) is widely regarded as a key indicator of hospital efficiency and quality of care [1–4]. Prolonged LOS is associated with higher rates of complications, healthcare costs and, possibly, hospital inefficiency. While several summary measures, transformations[5], and powerful analytic tools (generalised linear mixed models, machine learning) are available a reliable benchmark for LOS remains problematic[6–10]. An ideal LOS metric will include all hospitals, all patients, and all diagnoses.

By far the most common summary measure is the arithmetic mean (aLOS) - often reported without qualification, risk-adjustment, or precision[10]. This metric assumes that LOS data are normally distributed with a low coefficient of variation, where any variance at the patient level is exceeded by variance at the provider level. Since none of these assumptions holds true, a hospital with a larger proportion of complex patients or services will project the illusion of inefficiency. Furthermore, the numerator is often derived from the acute-care episode[3,11], rather than total LOS. Hospitals that code more ‘statistical separations’ will generate a smaller numerator, while the accompanying ‘statistical (re)admissions’ expand the denominator, and appear more efficient.

Although aLOS has its place (as a descriptive metric) it is misleading as a benchmark and comparator. For this purpose, an alternative metric is required. Since LOS data are right skewed (Figure E1, online Supplement), subject to truncation (exclusions), heterogeneity (time-dependent hazard), absorbing events (death), recurrent events (readmission), and autocorrelation[12–14], it would seem reasonable to consider LOS as a time-to-event metric. Time-to-event models adjust for such confounding, are readily available, and frequently employed in clinical research and survival analysis.

In this report, we employ time-to-event analytics to describe and compare hospital LOS. Our interest is risk-adjusted LOS and its relationship to the grand mean (state benchmark) and our primary purpose is methodological, rather than descriptive or comparative. A multiphase approach has been employed: Phase 1: data curation; Phase 2 and 3: stratification of risk; Phase 4: final model development; Phase 5 model validation. A review of all LOS metrics and available methods is beyond the scope of this report.

## METHODS

### Phase 1: Data curation

We accessed five-years of consecutive hospital records from the Victorian Admitted Episode Dataset (VAED[15]) for July 2019 through June 2024. We included all adult acute care separations and excluded paediatric separations (age<18 years), non-acute care (rehabilitation, geriatric assessment, palliative care, and mental health) separations, and records where the principal diagnosis (reason for admission) was missing. Each hospital was allocated to one of six peer group categories: public-tertiary referral, major metropolitan, major regional, public-other, private-tertiary or private-other.

Statistical separations/admissions during the same hospital stay were linked to the index episode to generate total hospital LOS, defined as the time difference between admission and discharge dates/times, rounded to the nearest hour and reported as fraction of days. In the VAED, each separation is allocated to one of three expected-LOS categories: same-day (no overnight stay), or over-night (maximum one night), or multi-day (more than one night) stay. Since the latter group is of primary interest and consumes the majority of hospital bed-days, the former (short-stay) groups were excluded. Summary statistics for LOS skewness and kurtosis and patients’ demographics were generated for each fiscal year. The presence of autocorrelation was identified using the Stata command *phtest*[16].

### Phase 2: Classification of admission diagnosis

The principal diagnosis (main reason for hospital admission) is derived from medical records by clinical coders using the Australian modification of the International Classification of Diseases version-10 (ICD10-AM) guided by national[11] and state[15] coding rules. Since any one of 14,000 ICD10-AM codes may be selected as the principal diagnosis, each was assigned to a Clinical Diagnosis Group (CDG) using a published algorithm, described in detail elsewhere[17]. Stratification of CDGs according to LOS was undertaken in Phase 3.

### Phase 3: LOS risk stratification of CDG

The following candidate variables were fitted to LOS using a mixed-effect parametric survival time model[16]: age (transformed to square root), sex (male), emergent admission category, age-care resident, hospital transfer (from similar or lower service level hospital), relationship status (single or not), an ethnicity surrogate (English language or otherwise), indigenous status, and an interaction term (emergency inter-hospital transfer) and reason for admission (principal diagnosis); hospital factors (activity volumes, service capability), year of separation, and in-hospital death were added as fixed effects. Using an iterative process, candidate variables were removed if they were either non-significant (p-value >0.157[18]), increased (consistent Akaike and Bayesian) information criteria[19], or increased the p-value of another more significant covariate. Similarly, parametric options[16] for hazard distribution were tested for their effect on information criteria.

The coefficients generated by the above (explanatory) model were employed to sort, aggregate, and rank each CDG (and constituent ICD10-AM codes) according to their risk for prolonged LOS. Note, these coefficients were not employed in the final (Phase 4) prediction model. The optimal number of ranked categories (and coefficient range for each) was identified from their influence on the model’s information criteria. In short, Phases-2 and -3 simply aggregated over ten thousand principal diagnoses (reason for admission) into less than a dozen stratified (LOS-risk) categories.

### Phase 4: Final model development

The study dataset was randomly split into development and validation (75:25) datasets and LOS was (arbitrarily) truncated at 30-days to exclude extreme outliers. A parametric survival time regression model based on optimal hazard distribution was, again, fitted to LOS in the development dataset and adjusted for the on-admission and patient-specific candidate variables identified in Phase 3. All post-admission and hospital-related factors (from the Phase 3 interim model) were excluded from the final model since the intention was to compare variation in LOS at the provider-level. The form of the estimator was,

*streg age male aged-care emergency transfer interaction rank-1 rank-2 … rank-i, distribution(hd) vce(cluster hospital)*

where *streg* is the Stata command for a survival-time estimator[16] fitted to the outcome LOS and adjusted for: *age* in years transformed to the square-root, male *sex, aged-care* resident, *emergency* admission status, hospital *transfer* to higher service level, relationship (single) status, an *interaction* term for unplanned inter-hospital transfer; and the ranked admission categories *rank-1* to *rank-i* from Phase 3; optimal hazard distribution (*hd*); with standard errors (*vce*) adjusted for clustering at the hospital level.

### Phase 5: Model validation

Model coefficients generated from the development model were applied to the validation dataset to generate a predicted LOS (pLOS) for each hospital separation. A standardised hospital LOS ratio (HSLR) was generated for each hospital in each year, according to the formula:

*HSLR = (total observed bed-days) / (total predicted bed-days)*

where a HSLR of unity indicates perfect calibration and a hospital within the jurisdictional benchmark; and HSLR >1.0 indicates that observed bed-days exceed predicted bed-days and a hospital that may be significantly different to the grand mean (benchmark).

Observed LOS data were tested for normality (*sktest*[16]), autocorrelation (*actest*[20]) and the predicted hazard tested for proportionality assumption (*phtest*)[16]. Model fit (in the validation dataset) was tested for calibration, classification and reclassification, and over-dispersion in the following manner. Calibration (goodness of fit) of the model was assessed using Cox-Snell residuals[16]. Classification was tested by recalibrating the model for each fiscal year (ending 30th June) and comparing HSLR values using a funnel plot with control limits set to ±2 and ±3 standard deviations (SD) and precision set to the predicted average number of occupied beds (as a surrogate for volume of hospital activity) with the user-written command *funnelcompar*[21]. The number of hospitals with a HSLR within the benchmark was ascertained.

Re-classification was quantified by comparing the number of hospitals with an outlier aLOS metric that were reclassified as HSLR inliers and vice-versa. Over-dispersion was quantified by the dispersion value (ϕ) and the standard deviation of the random effect (τ)[22] generated by the user-written command *funnelinst*[23]).

In summary, an ideal prediction model applied to a diverse group of hospitals with a uniformly high standard of care would be expected to produce low (Cox-Snell) residuals, reclassification of most, if not all, apparent aLOS outliers with fewer than 1% of HSLR values exceeding± 3SD; and minimal over-dispersion (ϕ value → unity, and τ → zero). Conditional and unconditional variance at the provider level was measured by the intra-class correlation coefficient (ICC)[24]. Sensitivity analyses of calibration and dispersion were undertaken for each hospital peer group and fiscal year.

This project was approved by the Department of Health and by the Eastern Health Human Research Ethics Committee (LR19/062), and the need for patient consent was waived. We employed Stata/MP™ v18.0 (2023, College Station, TX) statistical software. A TRIPOD[25] checklist is provided in Table E1 (online Supplement).

## RESULTS

In 2023 the estimated adult population of the state of Victoria was 5.28 million[26], served by 334 (public and private) acute health services. Over the five-year study period 219 (67%) hospitals reported 14.1million acute-care separations and 24.7 million total bed-days. After exclusions (Figure-1) there were 2.73 million (19%) separations and 15.53 million bed-days (63%) for analysis. Study population median LOS was 3.94 (interquartile range: 2.48-6.76) days and aLOS 5.68 ±4.98 days. A summary of population demographics is provided in Table 1.

**Figure 1.**
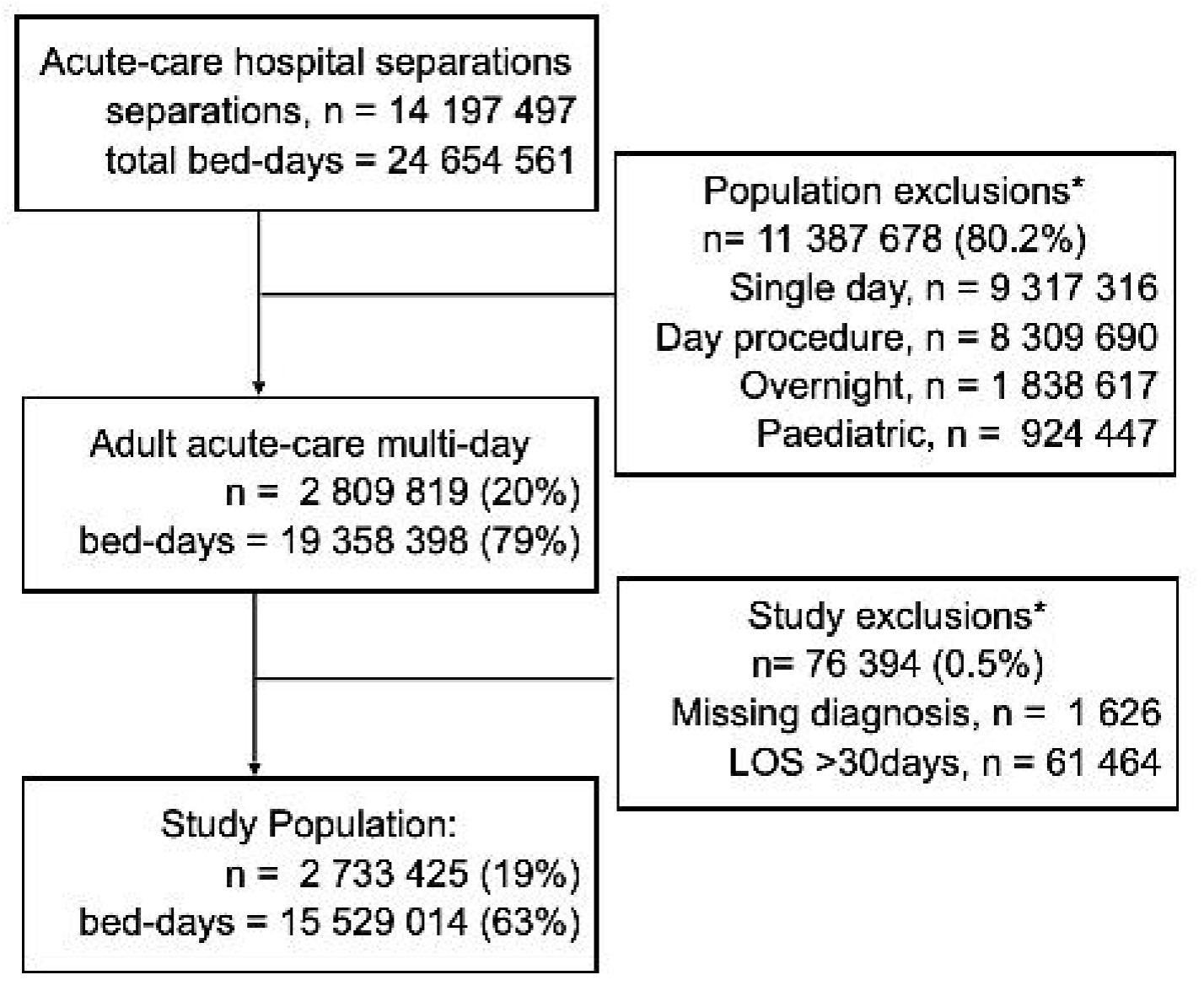
CONSORT diagram. LOS = length of hospital stay. Proportions refer to all acute-care separations. * Note, some records included in more than one exclusion category.

As expected, LOS was markedly right skewed (p<0.0001) (see Table E2 and Figure E1, Supplement) with autocorrelation (p<0.0001) and several observable (cyclical) patterns. Hospital discharge was delayed and LOS prolonged during the night shift, over weekends and public holidays and over the winter season.

A total of 12,145 (83.8% of the 14,487 available diagnosis codes in the Twelfth Edition[19] of the International Classification of Diseases and Health Related Problem, version 10, Australian modification [ICD10-AM]) were identified as principal diagnoses in the study dataset. These ICD10-AM codes were aggregated (during Phases-2 and -3) into nine ranked risk categories: Rank-1 includes any CDG with hazard ratio (HR) < 0.67; Rank-2 includes HR range of 0.68 - 0.75; Rank-3 HR = 0.76-0.82; Rank-4 HR = 0.83-1.00; Rank-5 HR = 1.01-1.20; Rank-6 HR = 1.21-1.33; Rank-7 HR = 1.34-1.50; Rank-8 HR = 1.51-1.80; and Rank-9 with HR >1.80.

Selected covariates (with a significant relationship to observed LOS) included: age, sex, emergent admission status, age-care resident, hospital transfer, relationship status, ethnicity surrogate, indigenous status, and nine ranked risk-categories for admission diagnosis. All but age were categorical variables. HSLR values demonstrated a distribution pattern approximating normality (Table E2 and Figure E2, Supplement). Cox-Snell residuals (Figure E3, Supplement) and information criteria favoured a lognormal hazard distribution. Model coefficients (Table E3) and worked examples (Table E4) are provided in the online supplement.

The study population was randomly divided into training (n= 2.05 million) and validation (n= 0.68 million) cohorts. The final model was fitted to LOS in the training dataset and the coefficients applied to the validation cohort to estimate pLOS. Model calibration in the validation dataset is represented by Cox-Snell residuals graph (Figure E3). Over 5-years, 219 hospitals provided LOS data for 940 hospital-years - twenty-five (small or new) hospitals did not report acute-care admissions in all five-years.

The majority (n= 572; 61%) of hospital aLOS values were outside ±3SD of the grand mean (state benchmark; Figure 2), even with adjustment for precision. Following risk-adjustment for patient-related factors present on arrival 936 (99.5%) HSLR values were found to be within ±3SD, and 98% within ±2SD of the state benchmark (Figure 3). The mean (±SD) dispersion metrics were, ϕ = 14.5 ±1.7, τ = 0.09 ±0.001 (Table E2, online Supplement). Variance at the hospital (ICC=0.025), peer group (ICC=0.008), and year level (ICC <0.0001) were small (Table E2).

**Figure 2.**
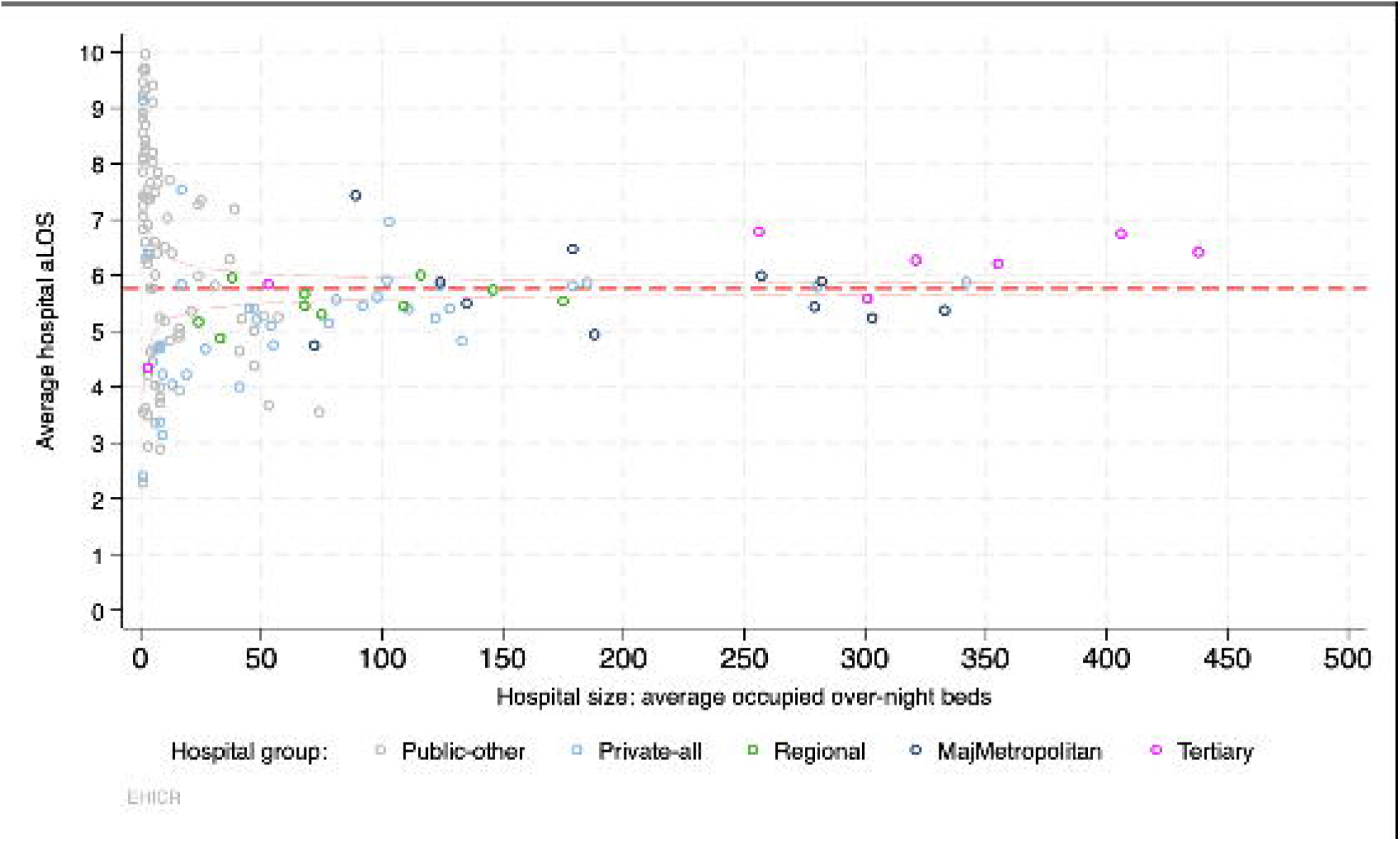
Example of funnel plot for hospitals aLOS, fiscal year 2023-24. Graph includes 568,083 separations and 187 hospitals but only 150 with ≥0.5 average overnight beds shown; red dashed line = weighted grand mean. See text for peer group definitions.

**Figure 3.**
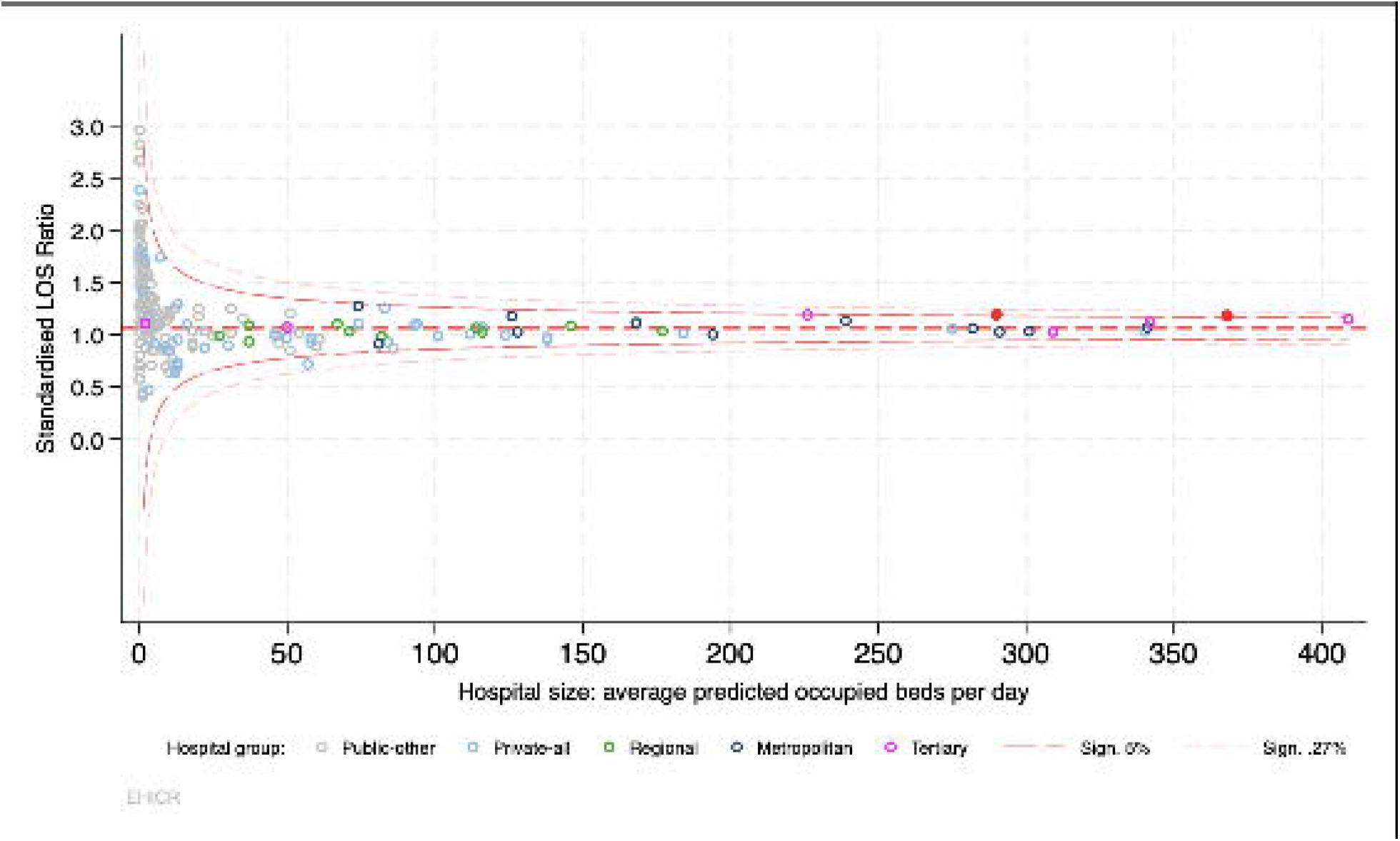
Example of funnel plot for hospital standardised LOS ratio (HSLR) for fiscal year 2023-24. Graph includes total 571,813 separations and 187 hospitals. Circles represent hospitals, red dots represent hospitals that exceed 95% confidence interval. No adjustment for over-dispersion included (see Table E1, Supplement). See text for peer group definitions.

Sensitivity analyses of hospital peer groups, fiscal years, and hospital survivors (excluding in-hospital deaths) revealed similar results to those of the total population. Nor did the addition of short-stay and day-procedure separations change the results.

## DISCUSSION

We analysed a comprehensive and contemporary population of 2.73 million acute-care adult separations from all adult acute care hospitals in Victoria, Australia, and confirmed the presence of substantial aLOS variation at the provider level, skewed distribution and autocorrelation. In contrast, the pLOS and HSLR values, derived from parsimonious time-to-event model and adjusted for patient characteristics present on admission, revealed much greater uniformity.

These findings are consistent with published reports[1–8]. While aLOS is a simple descriptor it is misleading as a performance comparator. In this study, two-thirds of observed aLOS values were outliers, yet, the standardised (HSLR) values resulted in reclassification of the majority as inliers. There are several plausible explanations for these contrasting findings. LOS revealed skewed distribution, autocorrelation and time-dependent (non-proportional) hazard distribution. Survival analysis adjusts for these confounders and may be a preferable choice over generalised linear models or standard Cox regression[27] models. These findings support the statistical validity of this approach and provide clinically plausible insights.

### Implications

The aLOS metric is a simple descriptor but a poor comparator. A standardised metric, such as the HSLR, based on a time-to-event (survival) analysis is also a simple ratio but a more robust comparator. These results are comparable across all hospitals and patient groups. The predominant cause of variance in aLOS at the provider level is variance at the patient level. The weight of current evidence suggests there is more uniformity between providers than previously appreciated. Whether or not the current model of healthcare produces the optimal LOS for most patients cannot be ascertained from these data. Further reduction in LOS for selected patients and diagnostic subgroups may be feasible, and warrant further detailed investigation. At the very least, these results support the conclusion of uniformity across providers.

### Strengths

This analysis has several strengths and, possibly, a number of unique features. Subjects were drawn from a contemporary jurisdictional population including all hospitals, all adult separations, and all acute-care admissions. The data source is the same administrative data common to Australian[11] and New Zealand health services. The methodology included adjustment for skewed data, competing risks, autocorrelation, and patient-related factors. We censored outcome at time of hospital, rather than episode, separation. We employed continuous time units (fraction of days) rather than a discrete interval (integer days) which, otherwise, generated an excess of tied (comparative) observations.

We employed a purpose-specific clinical classification system for admission diagnoses[17] rather than rely on the more common employed Diagnosis Related Groups or patient demographics. This approach has several advantages and enabled ranking of clinical risk and aggregation of over ten thousand ICD10-AM codes into nine ranked (diagnosis) categories. The resulting HSLR model is parsimonious yet demonstrates the crucial importance of adjusting for patient factors and standardising observed LOS. Finally, the (time-to-event survival) analytic model employed is common to most statistical software packages and easy to apply.

### Limitations

Clinical descriptors based on ICD10 codes lack granularity (clinical severity) and are dependent on the accuracy of coding and the quality of clinical documentation. Data were extracted from administrative datasets and not directly from the medical record. LOS remains complex and multi-dimensional. Despite favourable calibration and classification metrics our model remains insufficient for prediction of LOS in individuals or small groups, and over-dispersion was present even in this model. De-novo pLOS models applied to sub-populations of interest may be preferable to disaggregation and sensitivity analyses of an overall single model such as this.

Like most other LOS reports we excluded short-term (same-day and over-night) stay separations where further reduction in LOS is minimal and may be harmful (premature hospital discharge). Inclusion of these cases, however, did not change the results or conclusions. Nor did we exclude or adjust for (competing events such as) in-hospital death since these were infrequent (2.0%); but warrant separate analysis. The period of observation covered the entire duration of the SARS-CoV2 pandemic which may explain the rise in hospital mortality and prolongation of aLOS (p<0.001) in the first four years (Table-1). Further epidemiological analysis is now feasible with a benchmark for expected pLOS.

In conclusion, adult acute-care hospital separations consume a high proportion of hospital bed-days and observed aLOS varies widely between hospitals. The majority of this variation appears to reside at the patient-level, not the hospital level as is often assumed. Survival time models and the HSLR provide a more robust method for performance comparison. This methodology and these findings warrant validation in other jurisdictions and exploration in patient subgroups where avoidable excess LOS may still exist.

## Data Availability

All data produced are available online at the Victorian Agency for Health Information, Department of Health Victoria

https://vahi.freshdesk.com/support/home

## Acknowledgment

We thank the Department of Health Victoria for access to the original data, to all clinical coders and Health Information Managers who coded these records and the clinical staff who provide high-quality patient care, and Dr John Moran for statistical and clinical advice.

---

Table 1. Demographics of study subjects, for each fiscal year. SD = standard deviation, LOS = length of hospital stay; aLOS = mean LOS;

