## Supplement for "Hospital Standardised Length of Stay Ratio"

**ONLINE SUPPLEMENT**

**Manuscript title:** Hospital Standardised Length of Stay Ratio

Appendix 1: Table E1 TRIPOD statement for reporting of prediction models

Appendix 2: Table E2 LOS and Model Diagnostics

Appendix 3: Figure E1 Kernel density plot of LOS

Appendix 4: Figure E2. Kernel density plot of HSLR

Appendix 5: Figure E3. Cox-Snell goodness-of-fit plot

Appendix 6: Table E3 HSLR model variables and coefficients

Appendix 7: Table E4 Example of HSLR calculation

Appendix 8: Table E5 List of abbreviations

**Appendix 1**

TRIPOD statement for reporting of prediction modes

| | | TRIPOD Checklist : Hospital Standardised Length of Stay Ratio (HSLR). | Section, Paragraph |
|---|---|---|---|
| **Title and abstract** | | | |
| Title | 1 | Identify the study as developing and/or validating a multivariable prediction model, the target population, and the outcome to be predicted. | 1 |
| Abstract | 2 | Provide a summary of objectives, study design, setting, participants, sample size, predictors, outcome, statistical analysis, results, and conclusions. | 2 |
| **Introduction** | | | |
| Background and objectives | 3a | Explain the medical context (including whether diagnostic or prognostic) and rationale for developing or validating the multivariable prediction model, including references to existing models. | Introduction, Discussion |
| | 3b | Specify the objectives, including whether the study describes the development or validation of the model or both. | Introduction |
| **Methods** | | | |
| Source of data | 4a | Describe the study design or source of data (e.g., randomized trial, cohort, or registry data), separately for the development and validation data sets, if applicable. | Methods Data collection |
| | 4b | Specify the key study dates, including start of accrual; end of accrual; and, if applicable, end of follow-up. | Methods Phase 1, Results, para 1 |
| Participants | 5a | Specify key elements of the study setting (e.g., primary care, secondary care, general population) including number and location of centres. | Methods Phase 1 |
| | 5b | Describe eligibility criteria for participants. | Methods Phase 1 |
| | 5c | Give details of treatments received, if relevant. | Supplement |
| Outcome | 6a | Clearly define the outcome that is predicted by the prediction model, including how and when assessed. | Methods Phase 3 |
| | 6b | Report any actions to blind assessment of the outcome to be predicted. | n/a |
| Predictors | 7a | Clearly define all predictors used in developing or validating the multivariable prediction model, including how and when they were measured. | Methods Phase 3, Supplement |

| | | TRIPOD Checklist : Hospital Standardised Length of Stay Ratio (HSLR). | Section, Paragraph |
|---|---|---|---|
| | 7b | Report any actions to blind assessment of predictors for the outcome and other predictors. | n/a |
| Sample size | 8 | Explain how the study size was arrived at. | Results para 1. |
| Missing data | 9 | Describe how missing data were handled (e.g., complete-case analysis, single imputation, multiple imputation) with details of any imputation method. | 6 |
| Statistical analysis methods | 10 a | Describe how predictors were handled in the analyses. | Methods Phase 3 |
| | 10 b | Specify type of model, all model-building procedures (including any predictor selection), and method for internal validation. | Methods Phase 1, 2, & 3 |
| | 10 d | Specify all measures used to assess model performance and, if relevant, to compare multiple models. | Methods Phase 5 |
| Risk groups | 11 | Provide details on how risk groups were created, if done. | Methods Phase 3 |
| **Results** | | | |
| Participants | 13 a | Describe the flow of participants through the study, including the number of participants with and without the outcome and, if applicable, a summary of the follow-up time. A diagram may be helpful. | Figure 1 |
| | 13 b | Describe the characteristics of the participants (basic demographics, clinical features, available predictors), including the number of participants with missing data for predictors and outcome. | Tables 1, E2 |
| Model development | 14 a | Specify the number of participants and outcome events in each analysis. | Results, para 1, Figure 1. |
| | 14 b | If done, report the unadjusted association between each candidate predictor and outcome. | n/a |
| Model specification | 15 a | Present the full prediction model to allow predictions for individuals (i.e., all regression coefficients, and model intercept or baseline survival at a given time point). | Table E3 |
| | 15 b | Explain how to the use the prediction model. | Table E4 |
| Model performance | 16 | Report performance measures (with CIs) for the prediction model. | Results |
| **Discussion** | | | |
| Limitations | 18 | Discuss any limitations of the study (such as nonrepresentative sample, few events per predictor, missing data). | Discussion, Limitations |

| | | TRIPOD Checklist : Hospital Standardised Length of Stay Ratio (HSLR). | Section, Paragraph |
|---|---|---|---|
| Interpretation | 19 b | Give an overall interpretation of the results, considering objectives, limitations, and results from similar studies, and other relevant evidence. | Discussion, Strengths |
| Implications | 20 | Discuss the potential clinical use of the model and implications for future research. | Discussion, Implications |
| **Other information** | | | |
| Supplementary information | 21 | Provide information about the availability of supplementary resources, such as study protocol, Web calculator, and data sets. | Supplement |
| Funding | 22 | Give the source of funding and the role of the funders for the present study. | n/a |

**Appendix 2**

| Fiscal year | 2019-20 | 2020-21 | 2021-22 | 2022-23 | 2023-24 |
|---|---|---|---|---|---|
| **Separations, n** | 546 341 | 542 320 | 530 681 | 543 232 | 570 841 |
| **LOS, days mean (SD)** | 5.59 (4.86) | 5.63 (4.91) | 5.71 (4.96) | 5.79 (5.11) | 5.70 (5.02) |
| **ICC LOS, hospital level** | 0.017 | 0.023 | 0.018 | 0.025 | 0.022 |
| **LOS skewness** | 8.67 | 8.49 | 8.22 | 7.92 | 8.17 |
| **LOS sktest, chi-squared** | 295.5 | 295.5 | 285.2 | 290.4 | 285.2 |
| **HSLR skewness** | 1.30 | 1.18 | 1.26 | 1.18 | 1.50 |
| **HSLR sktest, chi-squared** | 39.1 | 31.0 | 34.3 | 32.1 | 47.4 |
| **HSLR dispersion value, $\phi$** | 13.2 | 13.9 | 14.4 | 15.9 | 14.3 |
| **HSLR random effect SD, $\tau$** | 0.09 | 0.09 | 0.09 | 0.10 | 0.09 |

Table E1 LOS and Model Diagnostics

LOS = duration of hospital stay; SD = standard deviation; ICC = intraclass correlation coefficient for LOS at provider level; sktest = joint test for skewness and kurtosis; HSLR = standardised hospital length of stay ratio.

**Appendix 3**

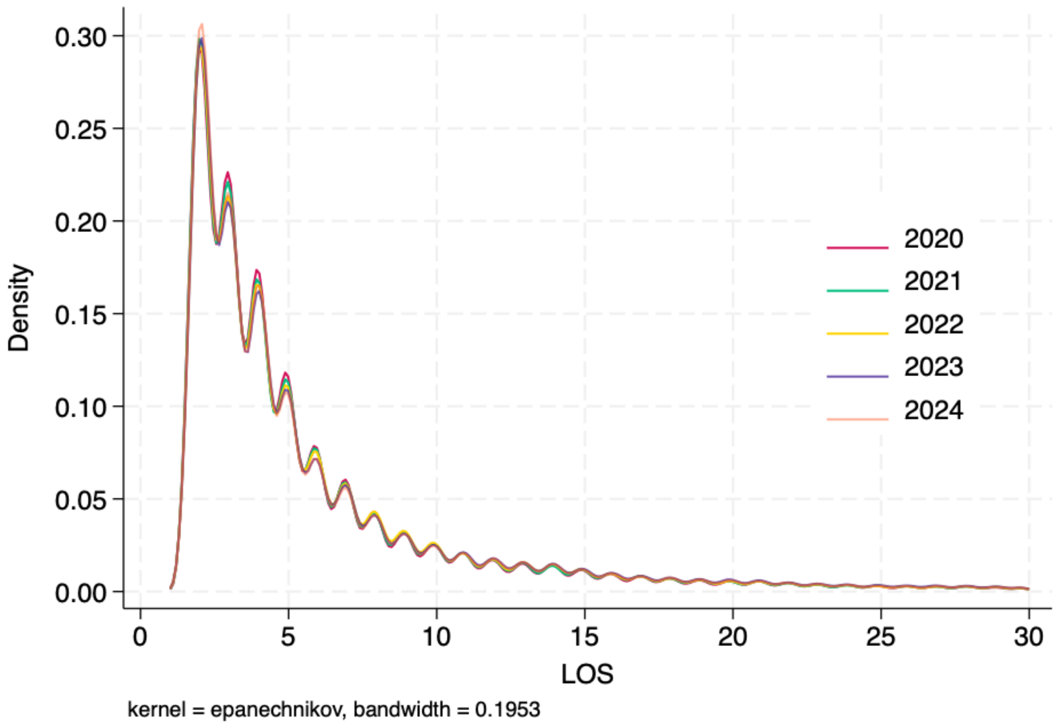


Figure E1. Kernel density plot of LOS for each fiscal year. Separations with LOS ≤1day excluded. Dataset truncated at 30-days.

**Appendix 4**

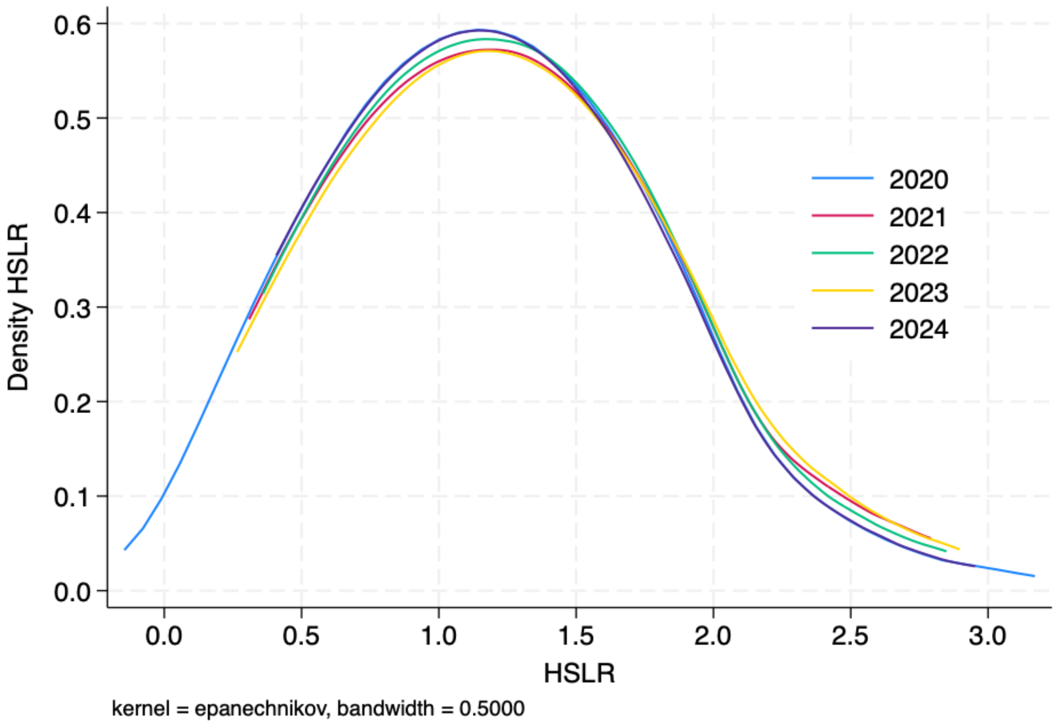


Figure E2. Kernel density plot of hospital standardised length of stay ratio (HSLR) for each fiscal year (ending 30th June).

**Appendix 5**

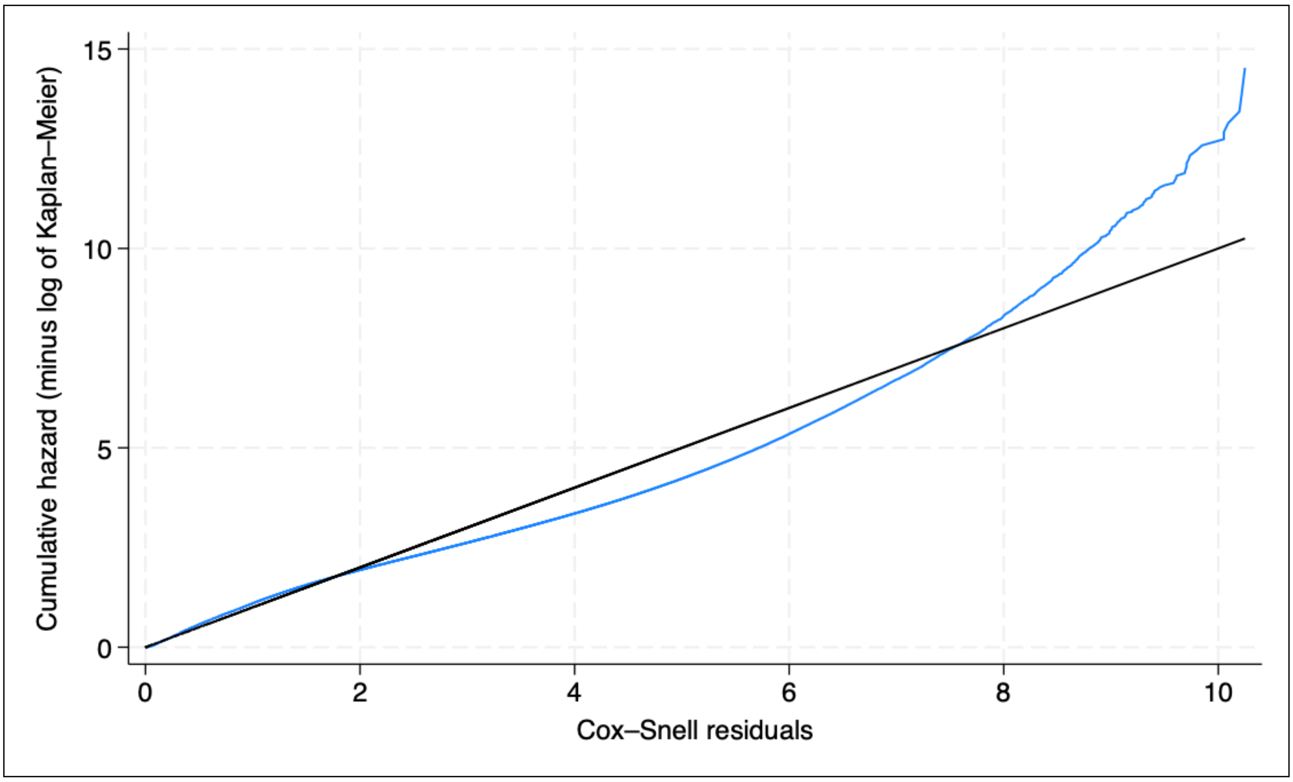


Figure E3. Cox-Snell goodness-of-fit plot for survival analysis with lognormal distribution, in the validation dataset (n=684,184). Solid black diagonal line = ideal model; solid blue line = predicted LOS.

**Appendix 6** LOS prediction model covariates, coefficients, and definitions

| **Variable** | **Coeff-icient** | **Description** |
|---|---|---|
| Age (years) | 0.116 | Age in years transformed to square root |
| Sex | -0.023 | Male=1; female=0, other=0 |
| Up-transfer from other hospital | 0.236 | Transfer from lower level of care =1 |
| Single | 0.078 | Relationship status no partner or spouse =1. |
| Non-english speaking | 0.050 | Primay language: english=0, other=1, unknown=0 |
| Unplanned admission | 0.039 | Non-elective admission=1; planned admission =0 |
| Indigenous status | 0.016 | Aboriginal or Torres Strait Islander =1 |
| Transfer admitted via hospital ED | -0.071 | ED admission transferred in from other hospital=1 |
| Aged care resident | -0.087 | Transfer from residential care facility = 1 |
| Constant | -0.449 | |
| **Diagnosis Category** | | **Clinical Diagnosis Groups**[17] |
| Category-1 | -0.480 | Cataract, dental disease, glaucoma, nontoxic goitre, prolapse, prostatomegaly, renal calculi. |
| Category-2 | -0.347 | Anaphylaxis, breast lump, chest pain, diverticulitis, endometriosis, ear disease, gastrointestinal polyp, hydronephrosis, mastoiditis, miscarriage/abortion, PV bleed, transient stroke, |
| Category-3 | -0.253 | Atrial fibrillation, appendicitis, bowel hernia, bradycardia or syncope, fractured jaw, gastritis, hyperparathyroidism, thyrotoxicosis, gastro-oesophageal reflux. |
| Category-4 | -0.109 | acute coronary syndrome, anorectal abscess, asthma, cerebrovascular occlusion, cholecystitis, chronic obstructive lung disease, gastrointestinal haemorrhage, infective gastroenteritis or colitis, migraine, minor head injury, pericarditis, pulmonary embolism, pyelonephritis, urological cancer. |

| Variable | Coeff-icient | Description |
|---|---|---|
| Category-5 | 0.069 | Acute kidney injury, anaemia/neutropenia, aspiration pneumonitis, bacterial and atypical pneumonia, autoimmune hepatitis, bowel obstruction, cardiac failure, cardiomyopathy, cellulitis, chronic liver disease, coagulopathy, colorectal cancer, connective tissue diseases, coronary artery disease, diabetic ketoacidosis, gastric or oesophageal cancer, immune deficiency, hypernatraemia. hypopituitarism, ischaemic stroke, pancreatic cancer, pleural effusion, pneumoconiosis, major chest or abdominal trauma, melanoma, metabolic acidosis, myelodysplasia, nephritis, toxic drug ingestion, vascular disease, viral hepatitis. |
| Category-6 | 0.234 | Amyloidosis, cardiac arrest, cholangitis, acute delirium, breast cancer, glomerulonephritis, hypothyroidism, hypovolaemia, metastatic cancer, hepatobiliary cancer, interstitial lung disease, lung cancer, non-Bcell lymphoma, mediastinal trauma, mesothelioma, severe traumatic brain injury, subdural haemorrhage, transplant rejection, valvular heart disease, viral pneumonia. |
| Category-7 | 0.329 | Acute peritonitis, alcoholic hepatitis, aortits, bone cancer, brain cancer, chronic kidney disease, chronic respiratory failure, gastrointestinal fistula or perforation, Guillain-Barre syndrome, haemorrhagic stroke, haemothorax, hepatic failure, hyperosmolar hyperglycaemia, meningitis, pelvic fracture, septic shock, wound infection |
| Category-8 | 0.455 | Aortic rupture, cardiogenic shock, dementia, femoral fracture, fungal infection, hip fraction, hyperosmolar coma, leukaemia, minor burns, myositis, opportunistic pneumonia, osteomyelitis, pressure injury, |
| Category-9 | 0.691 | Bone marrow transplant rejection, encephalitis, endocarditis, HIV/AIDS, necrotising fasciitis, opportunistic infections, severe burns, subarachnoid haemorrhage, tuberculosis. |

Table E3. Prediction model variables.

Note: each record is permitted any/all demographic variables but only one diagnosis Category derived from principal diagnosis ICD10-AM code. See reference #17 for ICD10-AM diagnosis codes specific to each Clinical Diagnosis Group.

**Appendix 7**

Example of LOS prediction and HSLR calculation.

Note: the information in the table below is based on fictitious cases as examples only.

| Variable | ß (model coefficient) | Patient 1 | Patient 2 | Patient 3 |
|---|---|---|---|---|
| **Reason for admission** | | Cardiac failure | Chronic kidney disease | Elective hernia repair |
| **Diagnosis rank** | | 5 | 7 | 3 |
| **Diagnosis coefficient** | | 0.069 | 0.329 | -0.253 |
| **Age (years)** | | 84 | 84 | 84 |
| **Age coefficient = ß x (√age)** | 0.116 | 1.061 | 1.061 | 1.061 |
| **Up-transfer from other hospital** | 0.236 | Yes | | Yes |
| **Single** | 0.078 | Yes | Yes | |
| **Non-english speaking** | 0.050 | Yes | Yes | |
| **Unplanned admission** | 0.039 | Yes | | |
| **Indigenous status** | 0.016 | Yes | | |
| **Admitted after transfer to ED** | -0.071 | | | |
| **Aged care resident** | -0.087 | | | Yes |
| **Model constant** | -0.449 | Yes | Yes | Yes |
| **Sum of coefficients** | ∑ ß | 1.100 | 1.069 | 0.594 |
| **Predicted LOS (days)** | $e^{\beta}$ | 3.004 | 2.912 | 1.812 |
| **Reported LOS (hours)** | | 84 | 65 | 60 |
| **LOS Ratio (Observed/Predicted)** | | 1.165 | 0.927 | 1.380 |

Three fictitious 84 year-old males with the same three clinical conditions (congestive cardiac failure, chronic kidney disease requiring ambulatory dialysis, and a large inguinal hernia) are admitted to hospital, but the reason for admission (principal diagnosis) differs for each.

**HSLR calculation**

Sum of observed LOS = 209 hours = 8.71 days; sum of predicted LOS = 7.73 days.

HSLR = ∑[observed LOS] / ∑[predicted LOS] = 1.13, 95% CI = 0.91-1.41.

Note that the HSLR value is unreliable for single patients or small groups (n<30) and should always be accompanied by statistical confidence interval (CI).

**Appendix 8**

**List of abbreviations**

AIC: Akaike information criteria

BIC: Bayesian information criterion

CDG: Clinical Diagnosis Group

CFR: case fatality rate

HSLR: hospital standardised length of stay ratio

HR: hazard ratio

ICC: intraclass correlation coefficient

ICD10-AM: International Classification of Diseases and Health Related Problems Tenth Edition, Australian Modification

LOS: length of hospital stay

pLOS: predicted length of hospital stay

SARS-CoV2: severe acute respiratory syndrome coronavirus-2

SD: standard deviation
